# Altered Saccades, Blink and Pupil Responses in Functional Motor Disorder: New insight into Neurobiological Mechanisms

**DOI:** 10.64898/2026.03.24.26349168

**Authors:** Katarína Šútorová, Heidi C. Riek, Isabell C. Pitigoi, Donald C. Brien, Brian C. Coe, Barbora Křupková, Lucia Nováková, Tomáš Sieger, Douglas P. Munoz, Tereza Serranová

## Abstract

**Background:** Functional motor disorder (FMD) is a common and disabling condition with incompletely understood pathophysiology. Eye-tracking offers a method to objectively examine cognitive and motor control processes and their underlying neural pathways. We aimed to quantify saccade, blink and pupil responses in FMD and healthy controls performing an interleaved pro-/anti-saccade task, and to investigate the relationships between oculomotor measures and motor and non-motor symptom severity.

**Methods:** We conducted video-based eye-tracking in 104 patients with clinically definite FMD and 115 age- and sex-matched healthy controls performing the saccade task. Patients completed questionnaires on depressive, pain-related, dissociative, non-motor somatic symptoms. Clinician-rated motor severity and centrally acting medication was recorded in FMD patients.

**Results:** Compared to controls, FMD patients showed increased anti-saccade error rates (p < 0.001), anticipatory saccades (p ≤ 0.003), altered blink distribution (p < 0.001), and reduced pupil dilation velocity (p < 0.001). However, reduced pupil dilation velocity was not significant in subsample of unmedicated patients. Higher anti-saccade error rates were significantly associated with depressive symptoms, pain severity, dissociative symptoms, non-motor somatic symptom burden, and motor severity (all p < 0.05).

**Conclusions:** We hypothesize that the altered saccade and blink responses result from altered processing in the frontal cortex and basal ganglia which provide critical input to brainstem oculomotor control areas in FMD. These results support neurobiological models proposing altered predictive and attentional processing underlying FMD. Association between oculomotor measures and symptom severity suggests that specific cognitive abnormalities may play a role in the pathophysiology of these symptoms in FMD.

**WHAT IS ALREADY KNOWN ON THIS TOPIC:** FMD is increasingly interpreted through predictive coding models suggesting abnormalities in predictions about motor and sensory states driven by abnormally focused attention. Yet the underlying neurobiology remains poorly defined. Empirical studies directly probing basic predictive processes in FMD are scarce, and implicit cognitive–motor interactions, particularly those involving motor learning and adaptation, have been insufficiently explored.

**WHAT THIS STUDY ADDS:** Only two previous studies have used eye-tracking in FMD, focusing mainly on diagnostic saccadic markers. Using time-series analyses of saccadic, blink, and pupillary data, we show abnormalities in inhibitory control, predictive processing, and implicit learning. Due to strong homology between human and primate neurophysiology and neuroimaging findings in oculomotor control, the findings can be linked to dysfunction within cortico–basal ganglia circuits.

**HOW THIS STUDY MIGHT AFFECT RESEARCH, PRACTICE OR POLICY:** Oculomotor abnormalities correlated with motor and non-motor symptom severity, indicating mechanistic relevance. The findings provide empirical support for predictive coding accounts and point to involvement of subcortical structures including projections from the frontal cortex to the basal ganglia. This highlights the value of studying cortico–basal ganglia circuits with implications for treatment and of developing oculomotor measures as potential biomarkers in FMD.

## INTRODUCTION

Functional motor disorder (FMD), also referred to as conversion or dissociative disorder, is a highly prevalent condition associated with significant disability.^1, 2^ Its clinical presentation is heterogeneous, involving a range of motor (i.e., tremor, weakness, and gait disturbances) and non-motor symptoms (i.e., sensory disturbances, pain, fatigue, cognitive difficulties, dissociative features, and mood symptoms), which altogether contribute to a significantly diminished quality of life.^3-5^

FMD is characterized by inconsistent motor symptoms,^4^ often improving when attention shifts away from the affected body part.^3^ Neuroimaging and neurophysiological studies reveal widespread underlying dysfunction across attentional, salience, self-agency, and multimodal integration systems.^4^ Current theoretical frameworks based on predictive coding models of brain function highlight the central role of aberrant motor and sensory predictions driven by abnormally focused attention.^6^ However, empirical support remains limited. There is a need for well-developed objective measures of attentional deficits, salience attribution, and predictive processing abnormalities, insight into how they affect both automatic and voluntary movements, and characterization of the underlying neural circuits.

Eye-tracking can provide important insights into cognitive and motor control, including attentional and salience processes and their underlying neural mechanisms.^7, 8^ Video-based eye-tracking provides non-invasive and accurate quantitative assessment of oculomotor responses for precise localization of cortical and subcortical dysfunction. Only two eye-tracking studies have investigated saccades in FMD.^9, 10^ However, these studies focusing on potential diagnostic markers did not include blink or pupil metrics and collected few trials.

Here we employ the interleaved pro-/anti-saccade task (IPAST), which can assess attentional processing by revealing abnormal sensory, motor, and cognitive processes.^8^ Participants must either look toward a peripheral visual stimulus (pro-saccade) or suppress this automated response and instead look in the opposite direction (anti-saccade). Pro-saccades are easy to perform; however, anti-saccades are more difficult, requiring engagement of top-down control processes. As suggested by abnormalities across multiple levels of motor control in FMD,^4^ we hypothesize that the frontal and basal ganglia signals required for saccade inhibition and voluntary saccade generation are impaired, producing more anticipatory saccades (i.e., guessing behavior) and anti-saccade direction errors.

We have recently described blink behavior in the IPAST; blinks are least likely to occur during highly task-relevant periods.^11^ We hypothesize that this behavioural pattern will be compromised in FMD.

We have previously identified a pupil dilation response in the IPAST preceding peripheral stimulus appearance, which likely reflects cognitive control.^12, 13^ We hypothesize this pupil dilation will be reduced in FMD due to impaired top-down control.

Altogether, these measures provide an objective evaluation of widespread brain function. We implement time-series analyses of saccadic, blink, and pupillary responses in FMD patients and healthy controls performing the IPAST to localize disease-related dysfunction. Additionally, we examine correlations between selected oculomotor and clinical measures. We hypothesize that a key marker of cognitive control obtained from the IPAST - specifically, anti-saccade error rate - will correlate to motor and non-motor symptom severity.

## METHODS

### Participants

The study included 104 patients with clinically definite FMD (17 males, 87 females) from the Movement Disorders Center at the General University Hospital in Prague recruited between November 2023 and November 2025. The study was approved by the Ethics committee of the General University Hospital in Prague (approval no. 37/19), and all participants gave written consent.

FMD was diagnosed according to Gupta and Lang criteria^3^ following a detailed clinical interview and examination by experienced movement disorders specialists (co-authors LN, TSe), based on positive signs of functional weakness and/or abnormal movements that were inconsistent and incongruent with known movement disorders. Exclusion criteria were the presence of other neurological comorbidities associated with abnormal brain structure or function (i.e., epilepsy, Parkinson’s disease) or major psychiatric disorders (i.e., definite or suspected schizophrenia-spectrum disorders, Tourette’s syndrome).

Age-/sex-matched healthy control participants (CTRL; n=115 (93 females)) were drawn from an extensive normative database recruited from greater Kingston, Canada.^11, 13, 14^ CTRL had no known neurological or psychiatric conditions, and normal or corrected-to-normal vision. Queen’s University Health Sciences and Affiliated Teaching Hospitals Research Ethics Board reviewed and approved this protocol (#6040314). We assembled the control group by first finding the next youngest and next oldest control participant of the same sex as each FMD patient. This resulted in two matches per FMD participant; we then eliminated duplicate matches (Supplementary Fig.1).

### Objective assessment of motor symptoms

Dominant (the most severe and/or frequent) and additional motor symptoms were classified by a clinician (co-authors LN, TSe) as functional weakness, tremor, dystonia, myoclonus, gait disorder, or speech disorder. The severity of the motor disorder was assessed using the Simplified FMD Rating Scale (S-FMDRS).^15^ Administration of psychotropic medication, including antidepressants, anticonvulsants, opioids, and dopamine receptor antagonists or agonists, was also recorded.

### Subjective assessment of motor and non-motor symptoms

Depressive symptoms were assessed using the Beck Depression Inventory (BDI-II).^16^

Pain was measured with the PainDetect Questionnaire for current, average, and maximum pain over the past 4 weeks. The composite score was calculated by averaging these subscales.^17^

Dissociation was assessed using the self-report Multiscale Dissociation Inventory (MDI),^18^ which measures six different types of dissociative responses (disengagement, depersonalization, derealization, emotional constriction/numbing, memory disturbance, and identity dissociation).

All patients evaluated their subjective motor and non-motor symptom severity according to the modified Patient-Health Questionnaire (PHQ-15).^19^ In addition to existing items addressing weakness, gait, and motor coordination, we added two items assessing tremor and jerks, and abnormal postures or spasms (F-PHQ). For analyses, only the non-motor subscale was considered.

### Recording Apparatus

Eye position and pupil area data were collected monocularly using a video-based eye tracker (Eyelink-1000 Plus monocular-arm; SR Research Ltd, Ottawa, Canada) with a sampling rate of 500 Hz. The experiment was set in a dark, windowless room, where participants were seated with their eyes at a fixed distance of 60 cm from the screen. The task was displayed on a 17-inch LCD monitor (1280x1024 pixels, 32-bit color, 60Hz refresh rate). Gaze position was calibrated and validated using a 9-point grid array. A maximum validation error of 1.5° and average validation error of < 1° were permitted for analyses.

### Interleaved Pro-/Anti-saccade Task (IPAST)

Participants completed the IPAST in two 120-trial blocks, each lasting approximately 7 minutes, with a short break to prevent fatigue. A total of 240 trials was chosen to ensure sufficient data and consistency with previous studies.^20^ Every trial began with a central fixation point (1000 ms duration, 0.5° diameter circle, 44cd/m^2^) on a black background, colored according to trial instruction (green = pro-saccade, red = anti-saccade). Following a 200 ms gap in which the screen was blank, a peripheral target (1000 ms duration, 0.5° diameter gray circle, 62cd/m^2^) appeared at 10° left or right of screen center. Participants were required to generate a pro-saccade toward this target or an anti-saccade in the opposite direction, depending on the trial instruction. Trial instruction (pro/anti) and target location (left/right) were both pseudo-randomly interleaved, and trials were separated by a 1000 ms intertrial interval consisting of a blank black screen.

### Data Analysis

We analyzed saccade, blink, and pupil behavior in FMD and compared it to healthy age- and sex-matched controls. Blink detection and trial categorization were done using an in-house auto-marking pipeline implemented in MATLAB (MathWorks Inc., Natick, MA, USA).^20^ Raw pupil area in pixels was measured continuously throughout the task. Further details of the time-series analysis of saccade, blink, and pupil responses are described in the Supplementary Methods.

#### Saccade analysis

IPAST saccade responses were categorized according to reaction time and correctness. Saccadic reaction time (SRT) was defined relative to stimulus onset, and invalid trials were excluded. Saccades were classified as anticipatory (−110 to 89 ms), express (90–139 ms), or regular latency (140–800 ms). Further details are provided in the Supplementary Methods.

#### Blink analysis

The start and end points of blinks were estimated by modelling noise in the pupil area data to define the boundaries of a period of eye-tracking data loss.^20^ Any data losses less than 50 ms or longer than 500 ms in duration were not considered viable blinks and were excluded.^11^ Blink probability, a robust and time-sensitive measure of blink behavior,^11^ was computed continuously across the entire trial.

#### Pupil analysis

Pupil analysis was completed using only pupil area data from the fixation and gap periods of the task, because the eyes were stationary at these times and the appearance of the central fixation point induces a standard pupil response: constriction followed by dilation prior to peripheral stimulus appearance.^12^ Trials containing blinks or saccades >2° in amplitude during these periods were excluded. We produced continuous curves of pupil size and velocity during the fixation period for correct pro-saccade and anti-saccade trials. Inter-individual variation was controlled for by dividing the curve for each trial by its baseline pupil diameter (measured as the mean pupil diameter from 150 to 200 ms after fixation point appearance) to yield pupil size responses as a proportion of baseline pupil area.

#### Correlation and regression analyses

We next investigated associations between anti-saccade direction error rate and motor and non-motor symptom severity (assessed using BDI, PainDetect, MDI, FPHQ, and S-FMRDS) using partial Pearson correlations adjusting with respect to the patients’ age, as anti-saccade direction error rates increased with age (on average, by 0.4% with each year of age, p=0.039)^14^. We also studied whether the anti-saccade direction error rate could be jointly explained in terms of BDI, PainDetect, MDI, FPHQ, S-FMRDS, age, and sex using a linear regression model. The correlation and regression analysis was conducted in a subset of 99 FMD patients (83 females, 16 males) due to missing data in the remaining cases. We selected anti-saccade direction error rate, as it is a frequently reported metric, providing information about response inhibition and attention.^8^ P-values were adjusted for multiple comparisons using Holm’s method. Correlations with corrected p-values of p < 0.05 were considered statistically significant.

## RESULTS

Cohort demographics are presented in Supplementary Fig. 1. No significant differences in age were observed between the FMD group (ages 18.0–67.3 years; mean=45.3, SD=11.9) and healthy controls (ages 18.0–67.4 years; mean=46.0, SD=14.4), (Welch’s t(215.26)=–1.22, p=0.223, Cohen’s d= –0.16).

Additionally, no statistically significant difference in sex distribution between the FMD group and the control group was found (χ²(1)=0.29, p=0.591, Cramér’s V=0.04). In the FMD group, the mean disease duration was 7.8 years (SD=6.9), and the mean S-FMDRS score was 12.2 (SD=7.2). Predominant and additional FMD phenotypes are shown in Supplementary Table 1.

### Saccade responses

There were significant differences in the distribution of SRTs between FMD patients and CTRL (Fig.1, Table 1). On pro-saccade trials, FMD patients exhibited an increased rate of anticipatory saccades (Fig.1A,B) and fewer correct pro-saccades at regular-latency SRTs than CTRL (Fig.1A). On anti-saccade trials, controls made more correct responses at regular latencies (Fig.1C), while FMD patients made more anticipatory saccades and direction errors (Fig.1D). Cluster-level statistics for all trial types are reported in Supplementary Table 2.

**Figure 1.**
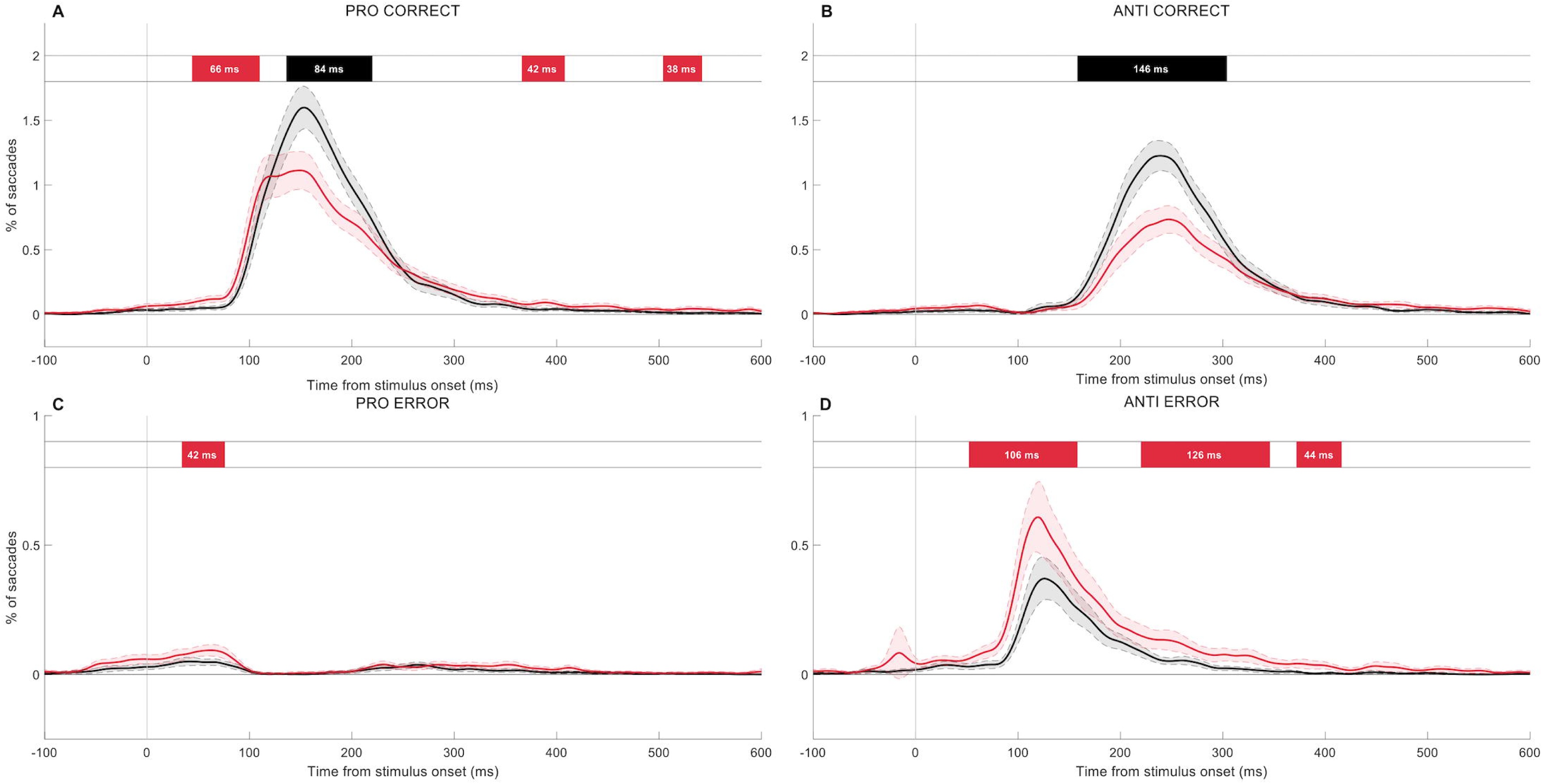
Distributions of saccadic reaction times (SRT: time from stimulus appearance to saccade initiation) for correct (**A,B**) and error (**C,D**) pro- and anti- saccade trials. Red = FMD; black = control. Shading represents 95% CI. Colored bars at top of graphs illustrate significant epochs (control > FMD = black; FMD > control = red).

### Blink responses

Blink probability was higher in FMD patients than CTRL on both pro- and anti-saccade trials (Fig. 2), with increased blinks occurring around fixation onset and the appearance of the peripheral stimulus. Cluster-level statistics for all trial types are reported in Supplementary Table 2.

**Figure 2.**
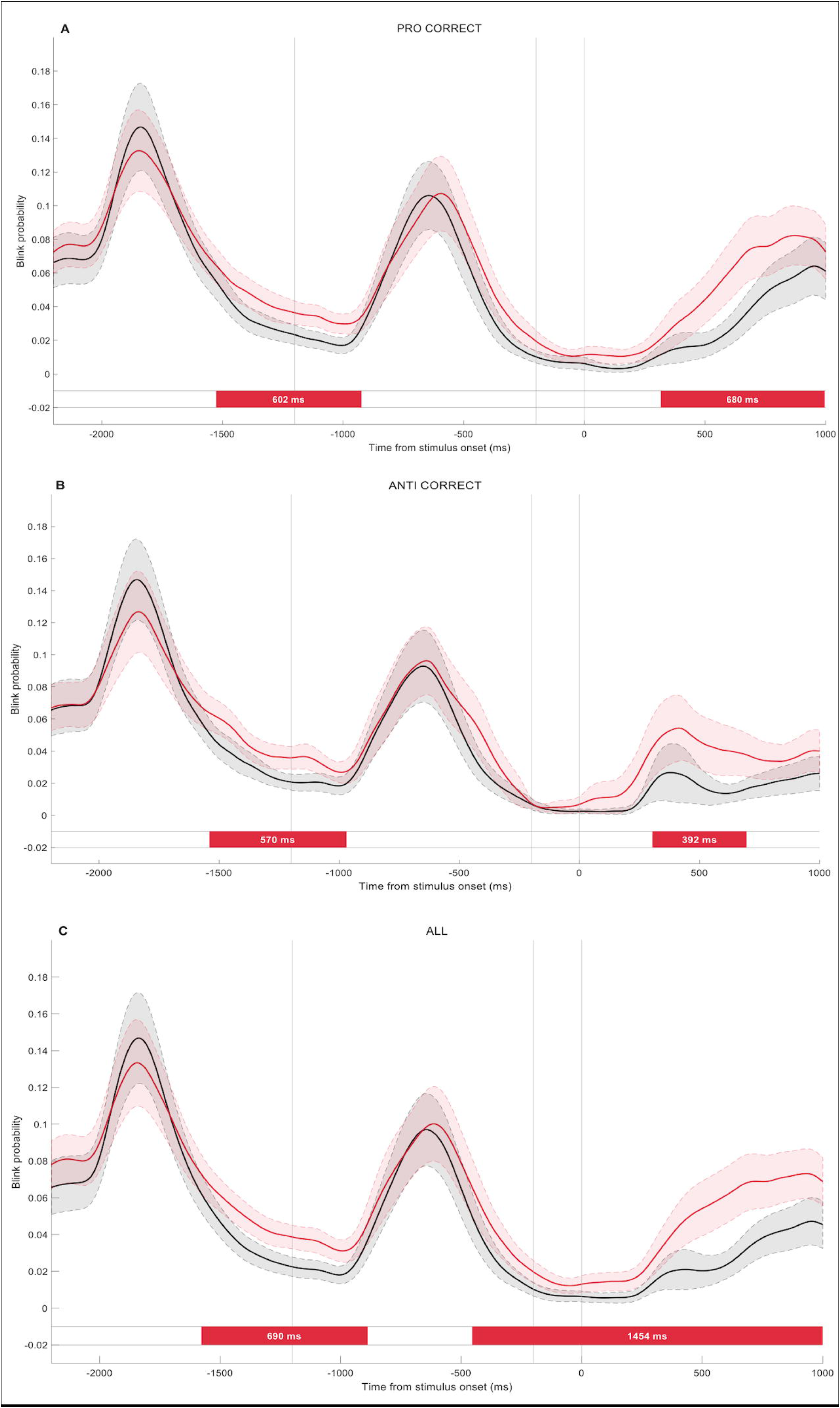
Blink responses in IPAST**. A**. Blink probability across the pro-saccade trials. **B**. Blink probability across the anti-saccade trials. **C**. Blink probability across all correct trials. Colored bars at bottom of graphs illustrate significant epochs (control > FMD = black; FMD > control = red).

### Pupil responses

On correct pro-saccade trials, FMD patients showed larger pupil size than CTRL (Fig. 3A) and slower pupil velocity (Fig. 3B) at early and later epochs, respectively, indicating a weaker constriction response and slower dilation. On correct anti-saccade trials, pupil size did not differ between groups (Fig. 3C), whereas FMD patients exhibited slower pupil velocity during dilation (Fig. 3D). Cluster-level statistics for all trial types are reported in Supplementary Table 2.

**Figure 3.**
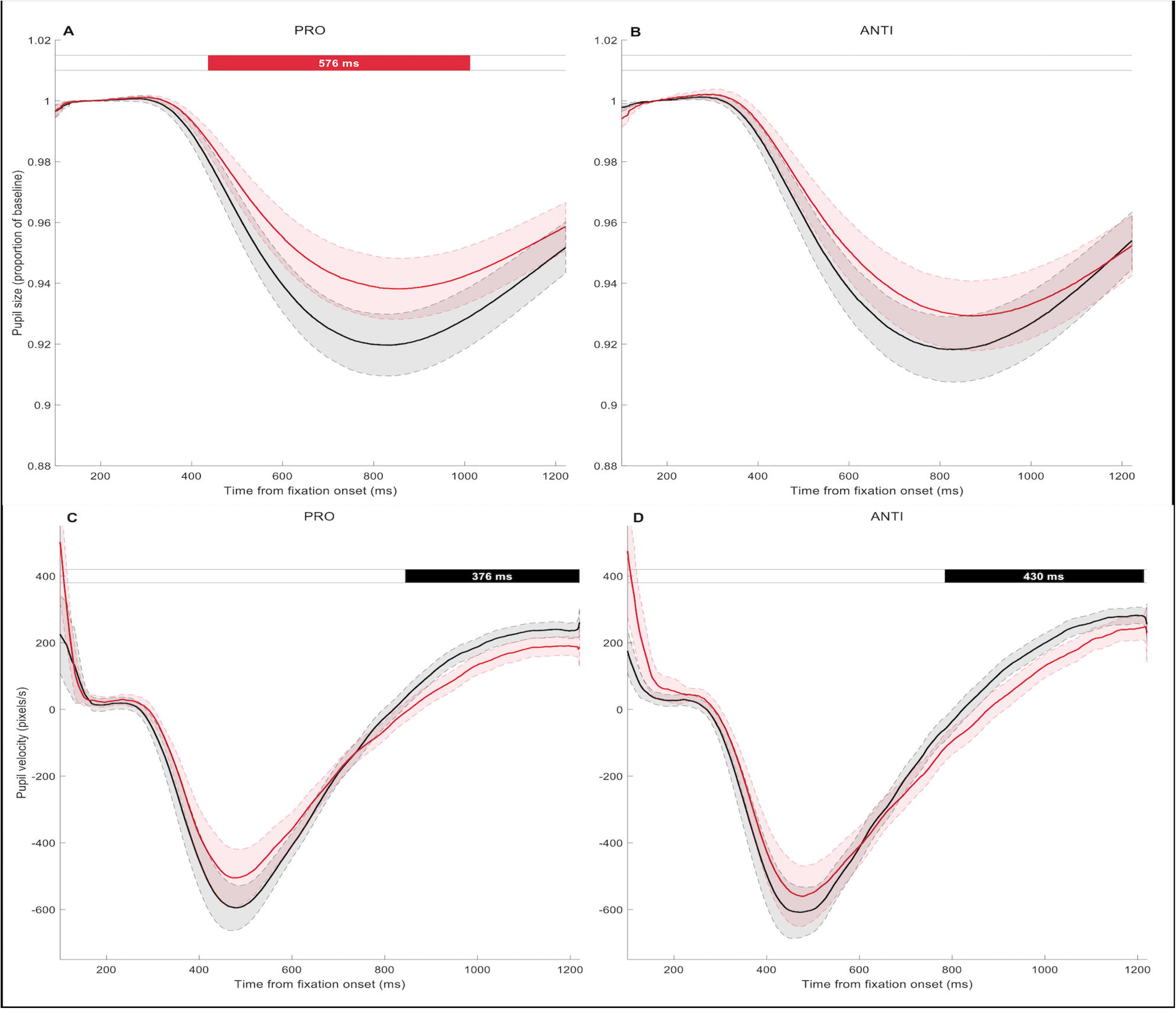
Pupil responses in IPAST. **A,B**. constriction followed by dilation during fixation of central fixation spot**. C,D**. Instantaneous pupil velocity aligned on fixation onset. Colored bars at top of graphs illustrate significant epochs (control > FMD = black; FMD > control = red).

### Correlation and regression analyses

These analyses were performed (Fig. 4) in a subset of 99 patients (ages 18.0–67.3 years, mean=44.8, SD=11.9). Higher anti-saccade error rates were significantly correlated with greater depressive symptom severity (r=0.26, p=0.011), higher pain severity (r=0.34, p=0.003), increased dissociative symptoms (r=0.29, p=0.008), greater non-motor somatic symptom burden (r=0.33, p=0.003), and greater motor severity (r=0.31, p=0.005). The regression model showed that the anti-saccade direction error rate could be jointly predicted by PainDetect (a unit increase on the PainDetect score, keeping other regressors fixed, was associated with anti-saccade direction error rate increase of 2.97%, p=0.016) and S-FMRDS (a unit increase of S-FMDRS was associated with anti-saccade direction error rate increase of 0.79%, p=0.016), and marginally by age (an increase of 1 year of age was associated with anti-saccade direction error rate increase of 0.34%, p=0.075). Other regressors, i.e. BDI, MDI, FPHQ, and sex did not have a significant effect on the anti-saccade direction error rate in the joint model (all p>0.14).

**Figure 4.**
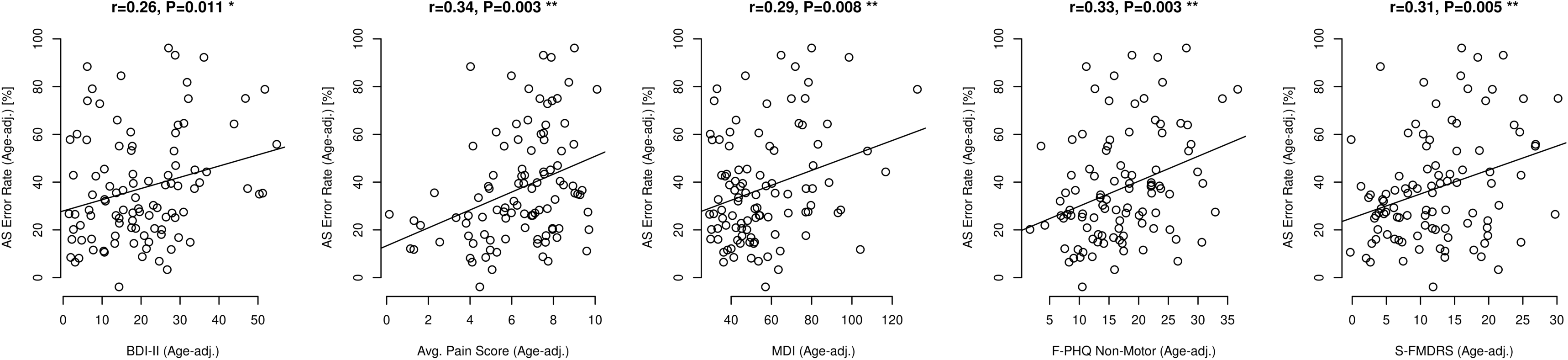
Partial correlations between clinical scores and anti-saccade error rate, adjusted with respect of age.

### Medication effects

Medication use was common among FMD patients, with many participants taking more than one type of medication: 50 participants (48%) were taking antidepressants, 36 (35%) anticonvulsants, 11 (11%) sedatives, 8 (8%) dopamine agonists, and 9 (9%) antipsychotics, while 31 FMD patients (30%) reported taking no medication acting on the central nervous system.

Between-group differences in saccadic, blink, and pupillary measures were further examined in a subset of unmedicated FMD patients (n=31 (28 females; ages 18.0 - 66.6 years; mean=45.6, SD=13.4)) and CTRL (n=115 (93 females; ages 18.0 - 67.4 years; mean=46.0, SD=14.4). Significant group differences were observed in anticipatory saccades, direction errors, and blink parameters, whereas no significant differences were detected in pupillary measures. Additional details are provided in Supplementary Material.

## DISCUSSION

Here we provided a detailed characterization of altered saccade, blink, and pupil behavior in a group of FMD patients with heterogeneous motor and non-motor manifestations. FMD patients showed increased rates of short-latency anticipatory saccades on both pro- and anti-saccade trials (Fig. 1), an increased frequency of anti-saccade direction errors (Fig. 1), and increased blink probability around the time of fixation point appearance (Fig. 2). Pupillometry revealed reduced pupillary dilation in FMD patients, a measure of cognitive processes reflecting motor preparation (Fig. 3), which may have been influenced by medication effects. Significant correlations between direction error rate and the severity of both motor and non-motor symptoms support the involvement of attentional dysfunction and impaired response inhibition mechanisms in FMD pathophysiology. Previous studies reported that FMD patients exhibit longer pro-saccade latencies, increased latency variability, and higher anti-saccade error rates compared to controls.^9, 10^ We replicated the increased frequency of anti-saccade direction errors in IPAST, for which the underlying neural mechanisms are well understood.

Saccades are triggered when premotor activity in the superior colliculus (SC) surpasses a threshold.^21^ To suppress short-latency pro-saccades on anti-saccade trials (direction error), the SC must be inhibited before appearance of the visual stimulus.^8, 22^ Frontal cortex and basal ganglia inputs to SC provide both this pre-stimulus inhibition and subsequent voluntary motor commands that generate a correct saccade in the opposite direction.^23^ Increased anti-saccade direction errors, as in FMD, reflect the impairment of these descending signals to the SC.^8^ We also found increased anticipatory saccades in FMD. These saccades, equally likely to be correct or incorrect in direction, reflect guessing behavior independent of sensory evidence.^20^ They are triggered when premotor activity in the SC exceeds the saccade threshold prior to the arrival of visual information.^24^

These results suggest impaired top-down input to the SC, in line with previous findings in FMD. Impaired intracortical inhibitory mechanisms have been demonstrated using TMS,^25^ plus abnormal subcortical inhibitory mechanisms under control of the frontal cortex and basal ganglia.^26^ Neuroimaging abnormalities have been observed across cortical regions involved in motor intention, planning, and preparation (e.g. abnormal connectivity of the dorsolateral prefrontal cortex and supplementary motor area), but also in motor execution and response inhibition (e.g., primary motor cortices, inferior frontal gyrus) and in interoception and multimodal integration.^4^ Neuroimaging also implies basal ganglia involvement, including volumetric changes, in FMD.^27^ Disrupted cortico-basal ganglia circuitry underlying response inhibition is additionally supported by poor Go/No-Go performance.^28^ Increased anticipatory saccades in FMD suggest a maladaptive trade-off: increased reliance on prediction and/or deficient inhibition produces excessive pre-stimulus SC activity at the cost of optimal oculomotor performance. In a probabilistic reasoning task, FMD patients formed beliefs after fewer cues than controls, indicating a “jumping to conclusions” bias.^29^ Increased anticipatory saccades could also reflect abnormal or excessive motor preparation, including reduced reliance on sensory evidence for decision-making.^30^ Such deficits could result from impaired attentional allocation or “top-down” cortical predictions overriding “bottom-up” inputs.^6^ Whether this disruption in hierarchical inference predisposes to FMD or reflects ongoing pathology remains unclear.

Recent evidence positions spontaneous blinking as a sensitive marker of cognitive and attentional control.^11, 31^ In healthy individuals, blink suppression is tightly time-locked to task-relevant events, particularly on correct anti-saccade trials where greater inhibitory control is required.^11^ In contrast, FMD patients showed increased blink probability surrounding fixation point appearance, when blinks are normally suppressed to facilitate processing of task-relevant visual stimuli.^11, 31^ This atypical behavior suggests impaired modulation of blinking in response to task goals, again likely reflecting disrupted interplay between attention, predictive control, motor inhibition, and implicit learning.

Impairments in anti-saccade errors, anticipatory saccades, and blink parameters were also observed in the unmedicated patient subgroup, suggesting that these abnormalities are intrinsic to FMD rather than attributable to medication effects.

Pupillometry revealed reduced pupil dilation velocity in FMD patients before stimulus onset on both pro-and anti-saccade trials (Fig. 3C, D). Normally, pre-stimulus dilation reflects preparatory activity for saccade initiation; larger pre-stimulus pupil size is associated with faster reaction times and lower error rates.^12^ Reduced pupil dilation velocity therefore suggests diminished preparatory engagement, consistent with reduced cognitive drive in FMD likely mediated via SC projections to the pupil.^32^

In our FMD cohort, most patients were taking centrally acting medications, often in combination, many of which can affect pupil responses. Analyses in the smaller unmedicated subgroup showed no significant differences, consistent with known medication effects. Beyond direct autonomic influences, these drugs may have sedative effects that impair cognition^33^ and alter sleep, further modulating pupillary dynamics. The effect of FMD itself cannot be excluded, as the number of medications correlated with both experienced and observed symptom severity, suggesting greater polypharmacy in more severe and cognitively affected patients. Although disentangling illness and treatment effects is therefore not possible, these results highlight the need for careful medication monitoring when interpreting pupillometric and other neurophysiological measures in FMD. However, the lack of differences may also reflect limited statistical power; moreover, our sample was insufficient to evaluate specific agents or combinations.

These results align with previous findings from neuropsychological assessments in FMD, which have identified attentional and working memory deficits.^34^ While typically FMD showed no significant changes in early stages of sensory or motor processing, evoked potential studies found differences at later stages of integration and abnormal correlates of preparatory and cognitive processes.^4, 35^ Alterations in correlates of perception-action integration processes were also found at the cortical level,^36^ where abnormal subcortical sensory information gating regulated by forebrain circuits involved in stimulus saliency was demonstrated using prepulse inhibition.^26^

Altogether, FMD abnormalities in the IPAST map well onto what is known in FMD and illustrate that FMD is a genuine disorder associated with multi-level brain dysfunction, including oculomotor control circuits.^4^ The predictive and attention-related abnormalities found in this study provide important support for current predictive coding models of FMD and extend current knowledge by identifying impaired inhibitory circuitry linked to specific behavioral outcomes.^6^

Importantly, we found that higher anti-saccade error rates correlated not only with clinician-rated motor symptom severity but also with self-reported depression, dissociation, pain intensity, and non-motor symptom severity. Anti-saccade error rate reflects attention, efficiency of voluntary oculomotor processes, and inhibitory control. In this study, poorer performance in the anti-saccade task was predicted by higher pain-related symptom severity and greater functional motor symptom severity, beyond the effect of age. This link supports the view of FMD as a multidimensional disorder and suggests that attentional dysregulation, abnormal saliency processing, and their interactions with motor control might play a role in generating and maintaining functional symptoms.^4, 5^ The observed association severity suggests that antisaccade performance may reflect a non-specific marker of cognitive-control vulnerability associated with broader overall disease severity or overall symptom load.

Oculomotor abnormalities in the IPAST have also been reported in various neurodegenerative and non-degenerative psychiatric and neurological conditions^37, 38^ and inhibitory control linked to cortico-basal-ganglia-thalamo-cortical circuits can be affected by many different pathologies.^39^ Therefore, comparative studies are needed to delineate the specific profiles of eye-tracking abnormalities in FMD and other functional somatic syndromes, such as fibromyalgia, irritable bowel syndrome, and mood disorders, which commonly co-occur with FMD and may share neural mechanisms.

This study has some limitations. First, the identified abnormalities are non-specific and should be understood as reflecting dysfunction within broader cortico–basal ganglia circuits rather than FMD-specific diagnostic markers. Second, heterogeneous medication use, including polypharmacy, reflects real-world practice but limits causal interpretation due to potential medication confounding, and may reduce the ability to detect subtle effects or disentangle the influence of individual medications.

This study revealed multiple saccadic, blink, and pupillary abnormalities in FMD, reflecting dysfunction in neural circuits involved in cognitive and motor control, response inhibition, and predictive processing. Association between motor and non-motor symptom severity and oculomotor markers underscore the relevance of these impairments in the pathophysiology of FMD. Video-oculography may thus serve as a sensitive tool to detect characteristic cognitive alterations and aid in developing objective diagnostic markers. Future work should clarify the specificity of these abnormalities relative to other neurological and functional disorders.

## CONFLICTS OF INTERETS

The authors declare that there are no conflicts of interest relevant to this work.

## DATA AVAILABILITY STATEMENT

The data analyzed in the present study are available upon reasonable request.

## RESEARCH ETHICS APPROVAL

The study complies with the Declaration of Helsinki’s ethical standards. The study was approved by the Ethics committee of the General University Hospital in Prague, approval no. 37/19. and all participants gave their written consent to take part in the study 53/23 Grant AZV VES 2024. The protocol was also approved by the Queen’s University Faculty of Health Sciences and Affiliated Teaching Hospitals Research Ethics Board (Protocol #6040314). We confirm that we have read the Journal’s position on issues involved in ethical publication and affirm that this work is consistent with those guidelines.

