## Supplementary Material for "Altered Saccades, Blink and Pupil Responses in Functional Motor Disorder: New insight into Neurobiological Mechanisms"

**SUPPLEMENTARY METHODS**

**Saccade analysis**

Saccadic reaction time (SRT) was calculated as the time when the first saccade was initiated relative to peripheral stimulus appearance. Trials with excessive tracking loss, very long SRT (>800 ms), saccades to random locations, failure to fixate the central fixation point, or no saccade response were removed. Saccades with SRT between 90–139 ms following stimulus appearance were categorized as express-latency saccades,^1^ while those with SRT between 140–800 ms were classified as regular-latency saccades. 90 ms is the minimum time required for visual signals to generate saccades; therefore, those with SRT 110 ms before to 89 ms after stimulus onset are deemed anticipatory saccades and indicate guessing behavior.

Express- and regular-latency saccades were further subdivided into correct responses and direction errors (away from stimulus for pro-saccades, towards for anti-saccades). Curves displaying the proportion of each response type as a function of SRT were constructed and used for further analysis (Fig. 1).

**Time-series analysis of saccade, blink and pupil responses**

Statistical analysis used the same workflow as a previous study which analyzed saccade, pupil, and blink data in time-series format.^2^ This method is called cluster-based permutation (CBP), an analysis method originally developed for EEG data^3^ but which has since been applied to continuous eye movement data^4, 5^ to detect differences between two groups or conditions over time. This method accounts for family-wise error rate and the autocorrelative nature of time-series data. In brief, we computed a two-tailed Wilcoxon rank-sum test (FMD vs. controls) at each time point, creating clusters of adjacent significant tests whose test statistics can be added to produce a cluster sum. We then completed 1000 permutations of the analysis, with each permutation consisting of shuffling an unlabeled version of the dataset with participants randomly sorted into two groups, re-performing the Wilcoxon analysis, and summing the test statistics within clusters of adjacent significant time points. The maximum cluster sum from each permutation was then stored to produce a null distribution after 1000 permutations, which became the basis for calculating p-values of clusters in the original labeled data.

Clusters with sum statistics outside the 2.5%–97.5% range of the null distribution were considered significant (i.e. p < 0.025). Clusters are displayed on horizontal tracks on the respective figures, colored according to which group was higher during the significant time window. Significantly different periods
(p < 0.025) were displayed with dark bars. These may be important to report due to the reduced ability of CBP analysis to detect smaller significant clusters because only the maximum cluster sum from each permutation is used to build the null distribution from which p-values are calculated.^3, 5^

**SUPPLEMENTARY RESULTS**

**Patient and control age distribution**

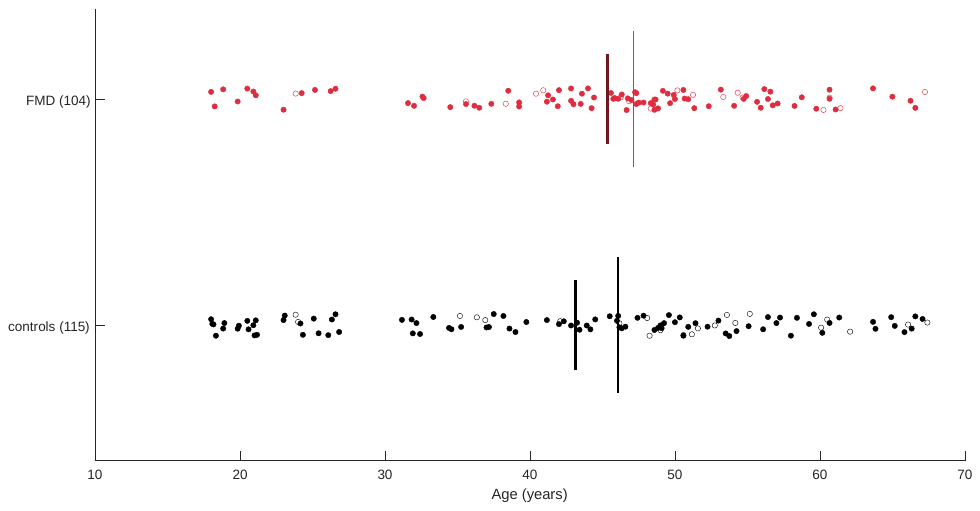
**Supplementary Fig. 1.** Distribution of participant ages for the FMD (red) and control (black) cohorts. Filled circles = females; empty circles = males. Tall thin vertical line = median age. Short thick vertical line = mean age.

**Predominant and additional FMD phenotypes**

| **Phenotype** | **Predominant n (%)** | **Additional n (%)** |
| --- | --- | --- |
| Weakness | 35 (34%) | 40 (38%) |
| Gait disorder | 30 (29%) | 40 (38%) |
| Tremor | 24 (23%) | 32 (31%) |
| Dystonia | 12 (12%) | 17 (16%) |
| Myoclonus | 3 (3%) | 7 (7%) |
| Speech and swallowing abnormalities | 0 (0%) | 9 (9%) |

**Supplementary Table 1.** Predominant and additional FMD phenotypes. Each participant could have one predominant phenotype and one or more additional phenotypes.

**Significant time clusters in saccade, blink and pupil measures between FMD patients and CTRL**

| **Effector** | **Trial Type** | **Cluster Sum** | **Cluster Begin ms** | **Cluster End ms** | **Cluster Length ms** | **Cluster P-Value** |
| --- | --- | --- | --- | --- | --- | --- |
| saccade | pro correct | -108.17 | 44 | 110 | 66 | 0.004 |
| saccade | pro correct | 163.80 | 136 | 220 | 84 | 0.001 |
| saccade | pro correct | -58.88 | 366 | 408 | 42 | 0.017 |
| saccade | pro correct | -60.09 | 504 | 542 | 38 | 0.016 |
| saccade | pro error | -67.21 | 34 | 76 | 42 | 0.001 |
| saccade | anti correct | 313.06 | 158 | 304 | 146 | < 0.001 |
| saccade | anti error | -176.77 | 52 | 158 | 106 | < 0.001 |
| saccade | anti error | -184.41 | 220 | 346 | 126 | < 0.001 |
| saccade | anti error | -63.53 | 372 | 416 | 44 | 0.011 |
| blink | all correct | -1403.12 | -1578 | -888 | 690 | 0.001 |
| blink | all correct | -2833.28 | -454 | 1000 | 1454 | < 0.001 |
| blink | pro correct | -983.68 | -1526 | -924 | 602 | < 0.001 |
| blink | pro correct | -1087.66 | 316 | 996 | 680 | < 0.001 |
| blink | anti correct | -804.54 | -1540 | -970 | 570 | < 0.001 |
| blink | anti correct | -573.95 | 304 | 696 | 392 | 0.006 |
| blink | anti error | -1201.22 | -1650 | -952 | 698 | < 0.001 |
| blink | anti error | -1297.30 | 238 | 1000 | 762 | < 0.001 |
| pupil size | pro correct | -791.98 | 436 | 1012 | 576 | 0.007 |
| pupil velocity | pro correct | 589.65 | 844 | 1220 | 376 | 0.001 |
| pupil velocity | anti correct | 669.81 | 784 | 1214 | 430 | < 0.001 |
| pupil dilation | pro correct | 293.30 | 80 | 300 | 220 | 0.009 |
| pupil dilation | anti correct | 418.18 | 14 | 300 | 286 | < 0.001 |

**Supplementary table 2.** Cluster-level statistics from cluster-based permutation analyses comparing FMD patients and CTRL (see Supplementary Methods). Cluster sum reflects the summed Wilcoxon test statistics within each temporal cluster. Cluster-level p values were derived from a null distribution constructed from 1000 permutations; clusters with p < 0.025 were considered significant. Pro correct / pro error indicate pro-saccade correct and error trials, anti correct / anti error indicate anti-saccade correct and error trials, and all correct refers to all correct trials regardless of trial type. Positive cluster sums indicate greater values in controls relative to FMD patients, whereas negative values indicate greater values in FMD patients. Times are reported in milliseconds relative to peripheral stimulus onset.

**Additional analyses in unmedicated FMD patients**

Between-group differences in saccadic, blink, and pupillary measures were further examined in a subset of unmedicated FMD patients (n = 31 (3 males, 28 females; ages 18.0 - 66.6 years; mean = 45.6, SD = 13.4)) and CTRL (n = 115 (22 males, 93 females; ages 18.0 - 67.4 years; mean = 46.0, SD = 14.4). Medicated FMD patients had significantly higher S-FMDRS scores than unmedicated FMD patients (Welch’s t(67.22) = 2.56, p = 0.006, Cohen’s d = 0.53).

**Saccade responses in unmedicated FMD patients**

There were significant differences in the distribution of SRTs between unmedicated FMD patients and CTRL (Supplementary Fig.2). On pro-saccade trials, unmedicated FMD patients exhibited an increased rate of anticipatory saccades (both correct and errors) (Supplementary Fig.2A, C) and fewer correct pro-saccades at early regular-latency SRTs than CTRL (Supplementary Fig.2 A). On anti-saccade trials, controls made more correct responses at regular latencies (Supplementary Fig.2B), while FMD patients made more anticipatory errors (Supplementary Fig.2D).

**Distributions of saccadic reaction times in FMD unmedicated group and CTRL**

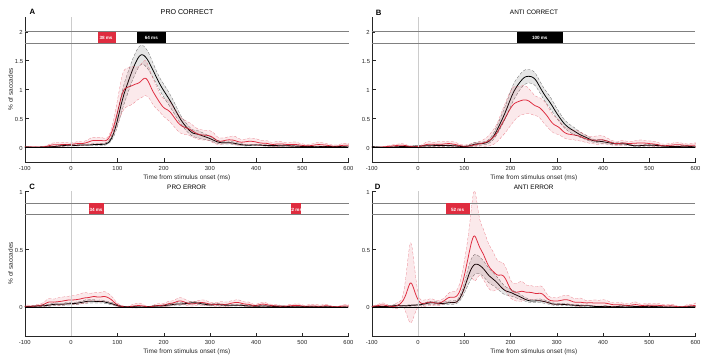

**Supplementary Fig. 2.** Distributions of saccadic reaction times in FMD unmedicated group (n=31) and CTRL (n=115), (SRT: time from stimulus appearance to saccade initiation) for correct (**A,B**) and error (**C,D**) pro- and anti- saccade trials. Red = FMD; black = control. Shading represents 95% CI. Colored bars at top of graphs illustrate significant epochs (control > FMD = black; FMD > control = red).

**Blink responses in FMD unmedicated group**

Blink probability was higher in unmedicated FMD patients than CTRL on both pro- and anti-saccade trials (Supplementary Fig. 3), with increased blinks occurring around fixation onset and the appearance of the peripheral stimulus.

**Blink responses in FMD unmedicated group and CTRL**

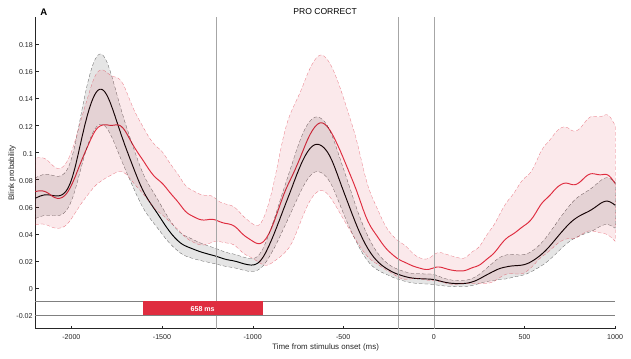

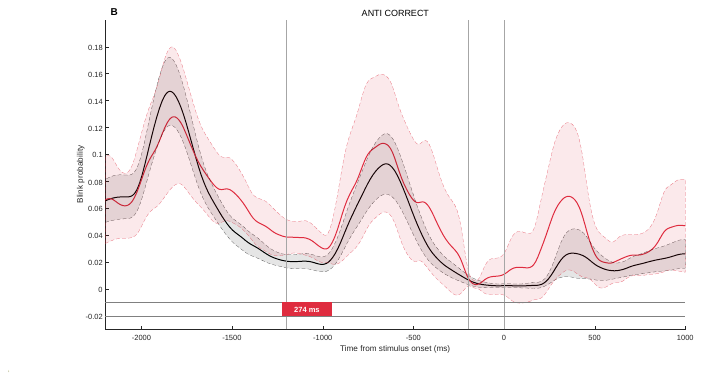

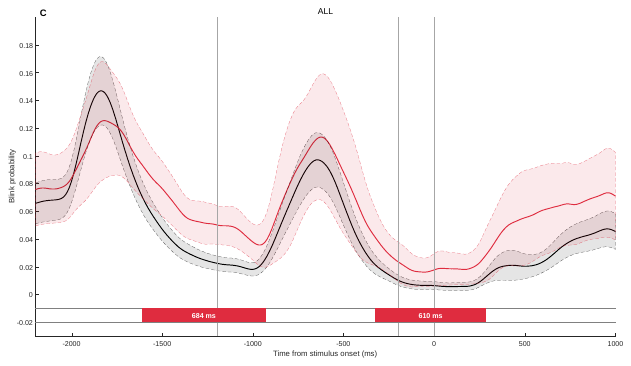

**Supplementary Fig. 3.** Blink responses in FMD unmedicated group (n=31) and CTRL (n=115). **A**. Blink probability across the pro-saccade correct trials. **B**. Blink probability across the anti-saccade correct trials. **C**. Blink probability across all correct trials. Colored bars at bottom of graphs illustrate significant epochs (control > FMD = black; FMD > control = red).

**Pupil responses in FMD unmedicated group**

On correct pro- and anti-saccade trials, pupil size and pupil velocity did not differ between groups (Supplementary Fig.4).

**Pupil responses in FMD unmedicated group and CTRL**

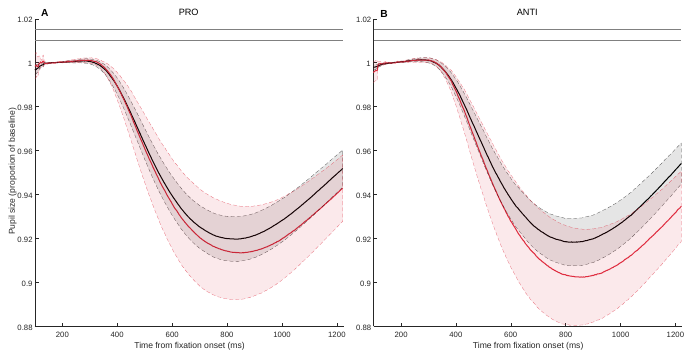

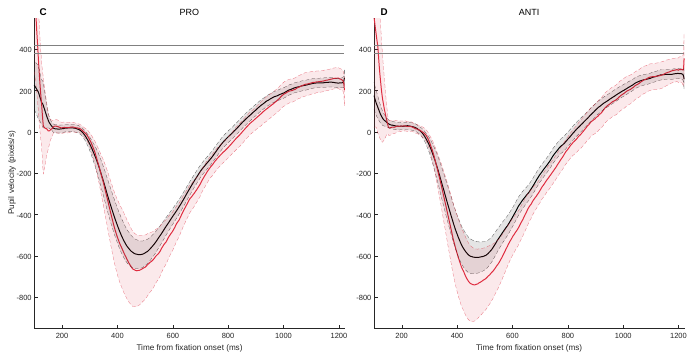

**Supplementary Fig. 4.** Pupil responses in FMD unmedicated group (n=28) and CTRL (n=115). **A,B**. Constriction followed by dilation during fixation of central fixation spot**. C,D**. Instantaneous pupil velocity aligned on fixation onset. Colored bars at top of graphs illustrate significant epochs (control > FMD = black; FMD > control = red).
